# HIV Testing Uptake and Late Diagnosis among Urban Population of Ethiopia: A Multilevel Modeling Approach

**DOI:** 10.1101/2021.07.16.21260636

**Authors:** Yimam Getaneh, Jemal Ayalew, Ebba Abate, Adamu Tayachew, Sileshi Lulseged, Yiming Shao

**Author notes:** **Corresponding Author:** Yimam Getaneh, Zhejiang University, School of Medicine, Hangzhou, China.

## Abstract

**Background:** Expanding HIV testing and early diagnosis requires a better understanding of determinants to uptake and late diagnosis. We investigated factors contributing to HIV testing uptake and late diagnosis.

**Objective:** The aim of this study was to determine level of HIV Testing Uptake and Late Diagnosis among Urban Population of Ethiopia

**Design:** We used data from the Ethiopian Population Based HIV Impact assessment in Ethiopia (EPHIA), conducted in 2017/18. EPHIA was a nationally-representative, cross-sectional and household-based study, conducted in urban Ethiopia which used a two-stage cluster sampling design with stratification into small and large urban areas. The EPHIA data were collected from 19,136 adults aged 15 to 64 years. For current study, we considered self-reports first diagnosis to estimate the testing uptake and also consider HIV LAg avidity Vs Viral Load Vs plasma Antiretroviral drug level algorithm to categorize the late diagnosis. In this analysis, a 2-level multilevel mixed-effect logistic regression model was employed using STATA v16. The effects of individual-level predictors were quantified by the estimates from the fixed-effect part of the model with a p-value less than 0.05 or 95% CI.

**Result:** By the year 2017/18, overall 29.4% of people living with HIV (PLHIV) were never tested in urban Ethiopia. Never tested male was significantly higher (32.4%; 95%: 31.0, 33.9) than females (26.4%; 95%: 25.3, 27.5) while it ranged from 28.3% to 47.8% among 45–54 and 15–24 years, respectively. By the same study period, 25.9% (95% CI: 21.7, 30.2) of PLHIV were lately diagnosed and male was significantly higher (36.8%; 95%: 28.0, 46.6) than females (20.8%; 95%: 17.3, 24.9). Late HIV diagnosis was also heterogeneous by various age group categories. Late diagnosis varies by region which ranged from 38.1% in the Gambella to 5.8% in Benishangul Gumuz. HIV testing uptake and late diagnosis was also significantly affected by alcohol use, income and educational status.

**Conclusion:** Ethiopia was behind the global target for HIV testing uptake. Late diagnosis of HIV constituted one-quarter of all testing HIV positive, and this was significantly higher among adult males. HIV testing offer and early diagnosis strategy need to focus on addressing geographically highly affected regions, and male and young adolescent age groups. The widowed population group, which is highly affected by HIV, and has low level of testing uptake, may require due attention by the HIV program. It is also suggested that HIV care and treatment programs provide a focus to the disadvantaged populations, especially those with limited education and low income.

## Background

In 2017, some 37 million people were living with human immunodeficiency virus (HIV), 70% of which were in sub-Saharan Africa (SSA)[1]. In Ethiopia, the HIV burden is high as in elsewhere in SSA, accounting for about 720,000 people living with HIV (PLHIV) and 27,104 newly diagnosed cases[2].

Voluntary counseling and testing (VCT) is considered the cornerstone for HIV prevention strategies and has been promoted as an entry point to prevention, care, and treatment services. Early HIV diagnosis and treatment access constitute one of the most effective ways to prevent the spread of the epidemic and protect the health of those living with the virus[3]. This helps to lower the viral load (VL), resulting in a dramatic reduction of the risk of morbidity and mortality among PLHIV, and in decreasing HIV transmission by greater than 90%[4], [5].

In order to mitigate the problem, Ethiopia adopted the global 90–90-90 HIV targets to be met by 2020. The targets include that by 2020, 90% of all PLHIV will know their HIV status, 90% of them will receive antiretroviral treatment (ART), and 90% these will have viral suppression[4][1], [6]. The achievement of these targets would contribute to achieving the longer-term strategic plan to eliminate the HIV/AIDS epidemic by 2030. A trend analysis suggests that Ethiopia could achieve around 79% of the first 90 target by 2020. Although through exerted toward the 90-90-90 goals held Ethiopia to reduce the incidence of PLHIV infection at the national and regional level, HIV still remains a major public health problem[7], [8].

The 2016 Ethiopian Demographic Health Survey (EDHS) showed that 57% of men and 60% of women were unaware of their HIV status among the 15–59 and 15–49 age groups, respectively[9]. In terms of residence, 35% of men and 32% of women were unaware of their HIV status in the urban population. The Ethiopian Population-based HIV Impact (EPHIA) has reported that the country achieved 79% of the first 90 target (awareness of their HIV status) among the urban population in 2018[6].

Many factors have been identified as barriers keeping people from knowing their HIV status, resulting in late presentation for HIV testing at the individual level. Being Male, 25-year old and above, being single, having education below the secondary level, low level of HIH and VCT knowledge, low perceived risk, stigma poor economic status, not listening to radio, not watching television, substance use, high depression level, having one sexual partner, and never got drunk were individual-level barriers[7], [10]–[12]. Similarly, factors contributing to late presentation for HIV diagnostics were being male, lack of knowledge about HIV/AIDS and ART, low perceived risk, older age, earlier year of presentation (before 2008), having two or more sexual partners, lower wealth index, HIV positive individuals with poor household social support, and fear of stigma[13]– [21].

Most of the studies cited above focused on individual-level risk factors. However, unobserved heterogeneity among environmental factors, like village and regional level factors, that may affect HIV testing behaviors are largely unknown. Environmental factors that are beyond individuals’ control may play a significant role in HIV transmission and acquisition. Understanding unobserved heterogeneity at the village and regional level may enhance the uptake of HIV testing and treatment. Hence, this study aimed to identify barriers to acquisition of knowledge by PLHIV about their HIV status, and delays in HIV diagnosis that explain both individual-level and unobserved regional characteristics using multilevel mixed-effect logistic regression model application to EPHIA data. This approach provides critical evidence on current barriers to testing which inform policies that encourage wider participation of the public in HIV testing.

### Objectives of the study

#### General Objective

- To determine the level of HIV Testing Uptake and Late Diagnosis among Urban Population of Ethiopia

#### Specific Objectives

- To determine the level of HIV testing uptake among people living with HIV in Ethiopia
- To determine the level of late HIV diagnosis among people living with HIV in Ethiopia
- Assess determinants of HIV testing uptake and late diagnosis among people living with HIV in Ethiopia

## Methods and materials

### Study Design

This analysis involved the EPHIA data, which was collected through a nationally-representative, cross-sectional and household-based survey conducted in urban Ethiopia from October 2017 to April 2018. The government of Ethiopia through the Ethiopian Public Health Institute (EPHI) of the Federal Ministry of Health conducted the survey with technical assistance from the United States Centers for Disease Control and Prevention (CDC) and ICAP at Columbia University, and in collaboration with local partners, including the Federal HIV/AIDS Prevention and Control Office (FHAPCO), the Central Statistical Agency (CSA) and the Ethiopian Public Health Association (EPHA). The EPHIA data were collected from 19,136 adults aged 15 to 64 years and 4,729 children aged 0 to 14 years. We included only the adult group for this analysis.

### Study Population and Sampling Procedures

EPHIA used a two-stage cluster sampling design with stratification into small and large urban areas. In the first stage 393 enumeration area (EAs)/clusters were selected using a probability proportional to size method based on EAs created by CSA for the 2007 Ethiopia Population and Housing Census, which included 17,339 EAs containing around three million households. The 393 EAs were further stratified by nine regional states and two city administrations: Tigray, Afar, Amhara, Oromia, Somali, Benishangul Gumuz, SNNPR, Gambella, Harari, Addis Ababa, and Dire Dawa. In second stage sampling, 30 households were randomly selected from each EA using an equal probability method, resulting in a total number of 11,810 households.

### Laboratory-based Biomarker Testing to identify late diagnosis

Twenty-one satellite laboratories for the study were established in existing health facility laboratories. One central reference laboratory (National HIV Reference Laboratory, EPHI, and Addis Ababa) was chosen for more specialized tests. At each satellite laboratory, trained laboratory technicians performed processing of whole blood specimens into plasma aliquots and DBS cards for storage at -20°C, testing for QA, and HIV confirmatory testing. For QA of the HIV rapid testing conducted in the field, the first 50 samples tested by each field tester and a random sample of 5.0% of specimens that tested HIV negative during HBTC, were retested in the laboratory using the national HIV rapid-testing algorithm. All specimens that tested HIV positive during HBTC, and those that had confirmed positive rapid test results during QA, underwent confirmatory testing using the Geenius HIV 1/2 Supplemental Assay (Bio-Rad, Hercules, California, United States). A positive Geenius result defined HIV-positive status for the survey. Central laboratory procedures included HIV VL testing, HIV DNA PCR for infant virological testing and for confirmation of status of those who self-reported an HIV-positive status but tested negative in HBTC, HIV recency testing, ARV drug resistance testing and long-term storage of samples at -80°C.

### Viral Load (VL) Testing

The HIV-1 VL (HIV RNA copies per mL) of confirmed HIV-positive participants was measured from plasma using the Roche (COBAS® AmpliPrep/COBAS® TaqMan® HIV-1 Test, Roche Diagnostics, Indianapolis, Indiana, United States) and from DBS using Abbott m2000 System (Abbott Molecular Inc., Chicago, Illinois, United States). Both instruments consist of two separate instruments, the sample preparation (Ampli prepap and m2000sp, which carries out automated extraction, purification, and preparation of HIV-1 RNA), and the Cobas Taqman-96 and m2000rt (which amplifies, detects, and measures the HIV-1 RNA load). In Cobas Taqman-96, 1 mL of plasma protocol was used, while the open-mode protocol for the Abbott Real Time HIV-1 assay was used to measure VL from DBS samples from children and adults with insufficient volume of plasma.

### HIV Recent Infection Testing Algorithm

To distinguish recent from long-term HIV infections, in order to estimate incidence, the survey used two different laboratory-based testing algorithms. Each algorithm employed a combination of assays: 1) HIV-1 LAg-Avidity EIA (Sedia Biosciences Corporation, Portland, Oregon, United States) and VL and 2) HIV-1 LAg Avidity EIA, VL, and ARV detection (Figure 1).

**Figure 1:**
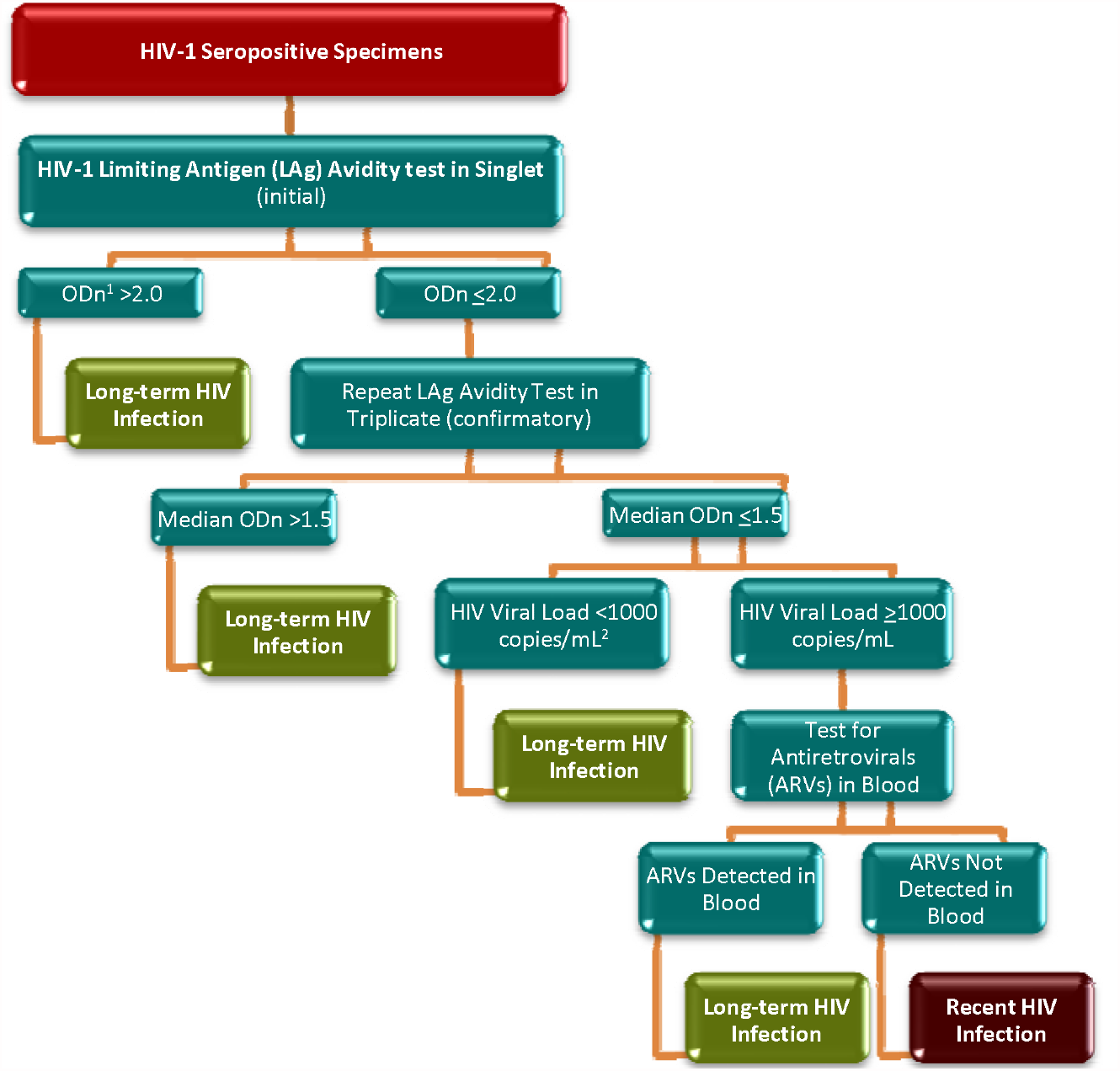
HIV-1 recent infection testing algorithm (LAg/VL/ARV algorithm) ^1^ODn: normalized optical density; ^2^mL: milliliter

Specimens with median normalized optical density (OD), ≤ 1.5 using LAg avidity testing was classified as potential recent infections, and their VL results were assessed. Specimens with VL < 1,000 copies/mL were classified as long-term infections, while those with VL ≥ 1,000 copies/mL were classified as recent infections. In the ARV-adjusted algorithm, specimens with VL ≥ 1,000 copies/mL and with detectable ARVs were classified as long-term infections. Specimens with VL ≥ 1,000 copies/mL and without detectable ARVs were classified as recent infections.

### Detection of Antiretrovirals

Qualitative screening, for detectable concentrations of ARVs, was conducted on DBS specimens from all HIV-positive adults by means of high-resolution liquid chromatography coupled with tandem mass spectrometry. The method used for ARV detection was a modified version of the methodology described by Koal et al. This qualitative assay was highly specific, as it separates the parent compound from the fragments, and highly sensitive, with a limit of detection of 0.02 μg/mL for each drug and a signal-to-noise ratio of at least 5:1 for all drugs. As detection of all ARVs in use at the time of the study was cost-prohibitive, three ARVs, efavirenz, lopinavir, and nevirapine, selected as markers for the most commonly prescribed first and second line regimens. Samples from participants who had suppressed viral loads and/or reported being on ART, but had no evidence of the first three compounds, were tested for nevirapine. These ARVs were also selected based on their relatively long half-lives, allowing for a longer period of detection following intake. ARV detection was performed by the Division of Clinical Pharmacology of the Department of Medicine at the University of Cape Town in South Africa.

### Study Variables

This analysis used two outcome (dependent) variables. The first outcome variable was HIV testing, a dichotomous variable coded as “1” when the study participant reported that he/she was never tested or received HIV test result (never tested) and “0” when he/she reported otherwise (ever tested and received test result). All study participants aged 15 to 64 years were asked whether they were tested for HIV or not, and we generated the first dependent variable based on the response. The second outcome variable was late HIV diagnosis among HIV positive participants. During the survey, the LAg avidity, VL, and ARV detection algorithm were employed for all PLHIV to distinguish recent from long-term infections. Among long-term infections, those never tested and received HIV results before the survey were considered as late diagnosis and coded as “1” and otherwise coded as “0” (all recent infections and long-term with HIV positive status before the survey).

The independent variables were categorized into two: individual-level and environmental variables. The individual-level variables were further categorized into two: socio-demographic and behavioral. The socio-demographic variables included age, gender, religion, wealth index, educational status, marital status and work status at the time of the survey. Behavior related variable considered was alcohol drinking, and environmental variables administrative region and size of the study area.

### Statistical Analysis

Data analysis was done using STATA 16.0 Descriptive analysis of the EPHIA data was done for each variable and the results weighted as recommended. Bivariate analysis with cross-tabulation and 95% confidence interval (CI) were employed to examine the association between the outcome variable (HIV testing and late diagnosis) and the selected predictor variables.

The EPHIA study followed a hierarchical data structure as the survey was employed a multistage stratified cluster sampling. Testing can be affected by unobserved regional-level factors. In Ethiopia, where there is a high level of variation in health policies and priorities by region due to political decentralization. Models used for analysis, therefore, have to account for associations among observations within clusters to make efficient and valid inferences. A multilevel logistic regression model was fitted to assess for regional variation in the two dependent variables and identify their association with the independent variables considered in the study.

In this analysis, a 2-level multilevel mixed-effect logistic regression model was employed. All the independent variables categorized as individual-level variables were considered level-1 variables and the region as level-2 variables. In the multilevel regression model, the effect of level-2 variable (regional) was quantified by intra-class correlation (ICC), the proportion of total variation in the response variable accounted for by the between-regional variation. The effects of individual-level predictors were quantified by the estimates from the fixed-effect part of the model with a p-value less than 0.05 or 95% CI that didn’t include unity.

The significance in improvement of multilevel model over the usual standard logistic model was checked by the chi-square test. A significance chi-square test result indicated that the multilevel model better fitted over the standard logistic model.

### Ethical considerations

The study protocol obtained ethical clearance from the institution review boards (IRBs) of the Ethiopian Public health Institute, Centers for Disease Control and Prevention (Atlanta), and Columbia University (New York). The EPHIA Data Analysis Advisory Committee at EPHI approved the data analysis. Study participants gave consent for this study in addition to the IRB approvals.

## Results

### HIV Testing

A total of 18,926 adults aged 15 to 64 years were included in this cross-sectional study to assess self-reported awareness of HIV testing in urban Ethiopia. Overall, 29.4% of them reported that they never got tested for HIV. The proportion never tested for HIV ranged from 18.8% in the Harari region to 20.3% in Afar to 32.6% in Oromia to 69.1% in the Somali region. This pattern was statistically significantly different across regions (Table1), which also shows the proportion never tested in small urban areas was 32.1%, and 26.8% among adults in large urban areas.

**Table 1:** Prevalence of self-reported HIV testing history by regions and size of urban areas prior to the survey in urban-based population of Ethiopia

| Variables | Ever tested | Never tested | Prevalence of never tested |  |  |
| --- | --- | --- | --- | --- | --- |
|  |  |  | % | 95% CI |  |
| Region |  |  |  |  |  |
| Tigray | 1029 | 314 | 22.5 | 19.6 | 25.6 |
| Afar | 641 | 161 | 20.3 | 16.5 | 24.8 |
| Amhara | 2157 | 818 | 26.6 | 23.8 | 29.7 |
| Oromia | 3006 | 1440 | 32.6 | 30.2 | 35.1 |
| Somali | 274 | 629 | 69.1 | 59.2 | 77.5 |
| Benishangul Gumuz | 567 | 227 | 28.6 | 24.2 | 33.4 |
| SNNPR | 1794 | 864 | 32.4 | 30.0 | 35.0 |
| Gambella | 565 | 192 | 25.0 | 20.0 | 30.9 |
| Harari | 552 | 138 | 18.8 | 14.1 | 24.5 |
| Addis Ababa | 2015 | 753 | 25.2 | 23.1 | 27.3 |
| Dire Dawa | 565 | 204 | 25.8 | 21.1 | 31.0 |
| Area |  |  |  |  |  |
| Small | 6155 | 2988 | 32.1 | 30.4 | 33.8 |
| Large | 7031 | 2752 | 26.8 | 25.0 | 28.6 |
| Total | 13186 | 5740 | 29.4 | 28.3 | 30.5 |

As shown in Table 2 the proportion of never tested among adult males was significantly higher, 32.4% (95%: 31.0, 33.9) than females, 6.4% (95%: 25.3, 27.5). Nearly on-half of those who were never married, 49.5% (95% CI: 47.7, 51.3) were never tested. The proportion never tested was more than double among the married or cohabiting, 17.3%) 95% CI: 16.1, 18.6, and the divorced or separated (19.4%; 95% CI: 17.3, 21.8), and was almost twice among widowed, 28.3% (95% CI: 24.6, 32.2).

**Table 2:**
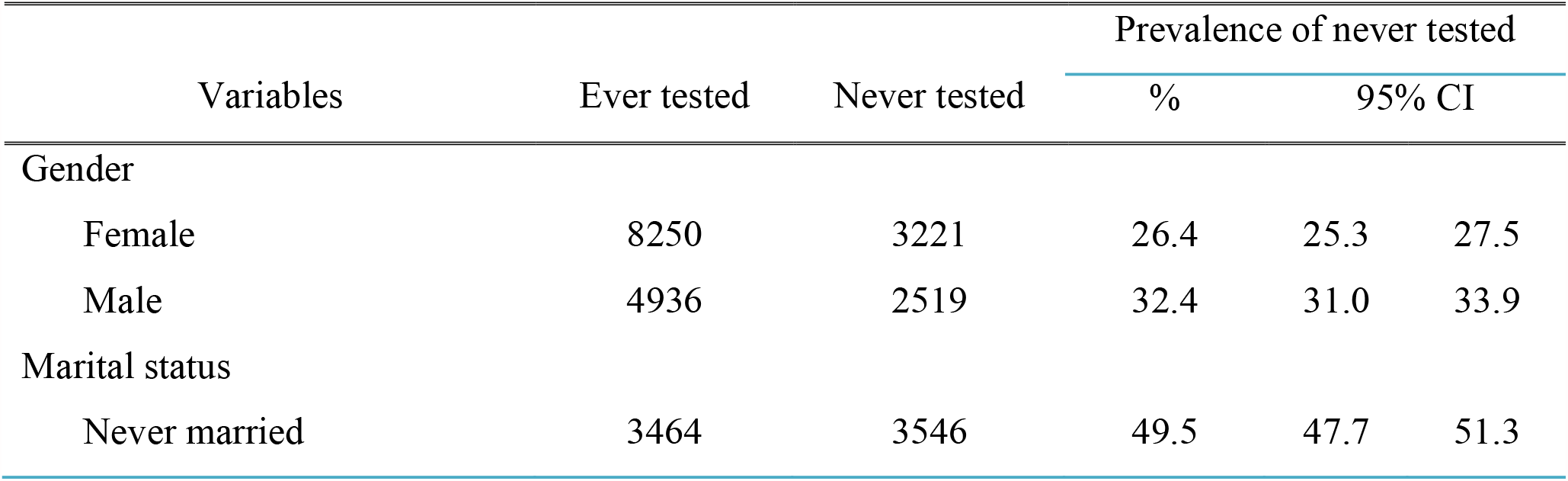
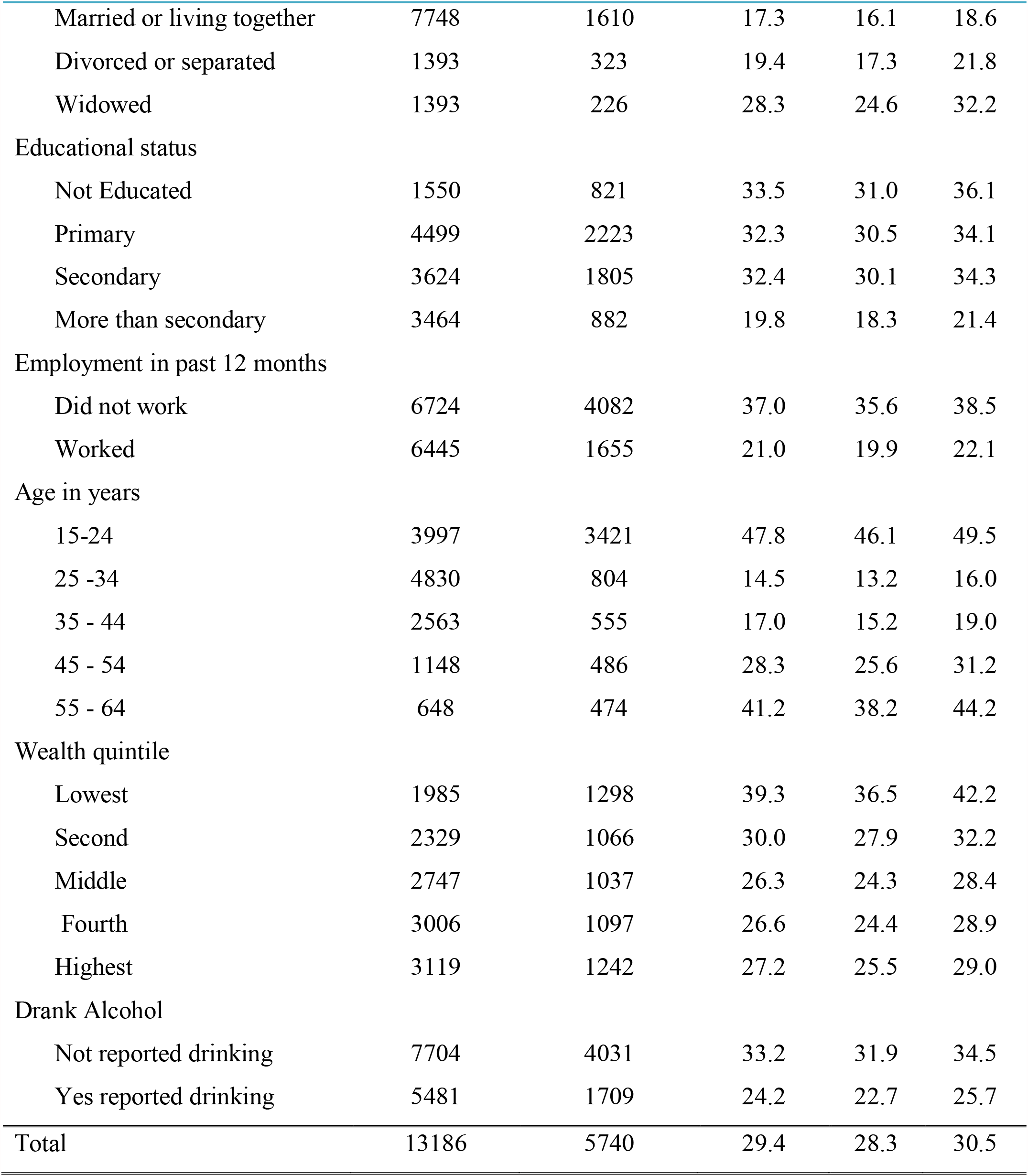
Prevalence of self-reported HIV testing history by some demographic and behavioral characteristics prior to the survey in urban-based population of Ethiopia (the significance test was mentioned using non-overlapping 95% CIs)

| Variables | Ever tested | Never tested | Prevalence of never tested |  |  |
| --- | --- | --- | --- | --- | --- |
|  |  |  | % | 95% CI |  |
| Gender |  |  |  |  |  |
| Female | 8250 | 3221 | 26.4 | 25.3 | 27.5 |
| Male | 4936 | 2519 | 32.4 | 31.0 | 33.9 |
| Marital status |  |  |  |  |  |
| Never married | 3464 | 3546 | 49.5 | 47.7 | 51.3 |
| Married or living together | 7748 | 1610 | 17.3 | 16.1 | 18.6 |
| Divorced or separated | 1393 | 323 | 19.4 | 17.3 | 21.8 |
| Widowed | 1393 | 226 | 28.3 | 24.6 | 32.2 |
| Educational status |  |  |  |  |  |
| Not Educated | 1550 | 821 | 33.5 | 31.0 | 36.1 |
| Primary | 4499 | 2223 | 32.3 | 30.5 | 34.1 |
| Secondary | 3624 | 1805 | 32.4 | 30.1 | 34.3 |
| More than secondary | 3464 | 882 | 19.8 | 18.3 | 21.4 |
| Employment in past 12 months |  |  |  |  |  |
| Did not work | 6724 | 4082 | 37.0 | 35.6 | 38.5 |
| Worked | 6445 | 1655 | 21.0 | 19.9 | 22.1 |
| Age in years |  |  |  |  |  |
| 15-24 | 3997 | 3421 | 47.8 | 46.1 | 49.5 |
| 25 -34 | 4830 | 804 | 14.5 | 13.2 | 16.0 |
| 35 - 44 | 2563 | 555 | 17.0 | 15.2 | 19.0 |
| 45 - 54 | 1148 | 486 | 28.3 | 25.6 | 31.2 |
| 55 - 64 | 648 | 474 | 41.2 | 38.2 | 44.2 |
| Wealth quintile |  |  |  |  |  |
| Lowest | 1985 | 1298 | 39.3 | 36.5 | 42.2 |
| Second | 2329 | 1066 | 30.0 | 27.9 | 32.2 |
| Middle | 2747 | 1037 | 26.3 | 24.3 | 28.4 |
| Fourth | 3006 | 1097 | 26.6 | 24.4 | 28.9 |
| Highest | 3119 | 1242 | 27.2 | 25.5 | 29.0 |
| Drank Alcohol |  |  |  |  |  |
| Not reported drinking | 7704 | 4031 | 33.2 | 31.9 | 34.5 |
| Yes reported drinking | 5481 | 1709 | 24.2 | 22.7 | 25.7 |
| Total | 13186 | 5740 | 29.4 | 28.3 | 30.5 |

There were significant variations among the never tested by education status, and the proportion of those never tested for HIV was significantly higher among adults with no education, primary education, or secondary education, 33.5% (32.3%, 32.4%,), respectively, compared to those with more than secondary school education (19.8%). Similarly, the proportion of adults who were reportedly never tested for HIV ranged from 14.5% in the 15–24 years category to 28.3% in 45–54 years to 41.2% in 55–64 years to 47.8% in 15–24 years. This pattern was significantly different across the age categories with non-overlapping 95% CIs.

In terms of wealth quintile, 39.3% of the adults who were in the lowest quintile had never tested for HIV, significantly more than from respondents who were in the second, third, fourth, and highest quintiles (30.0%, 26.3%, 26.6%, and 27.2%, respectively) with non-overlapping 95% CIs. Similarly, the proportion of never tested for HIV among adults who were not employed (during the last 12 months) preceding the survey was 37.0%, significantly more than almost twice among employed (during last 12 months) (21.0%) with the non-overlapping 95% CIs. Among the behavioral characteristics, the proportion of never tested for HIV was significantly higher in adults who didn’t report drinking alcohol, 33.2% (95% CI: 31.9, 34.5) compared with adult who used alcohol, 24.2% (95 % CI: 22.7, 25.7).

### Late HIV Testing

Among adults aged 15 to 64 years tested for HIV in EPHIA, 614 (3.0%) adults tested positive. A total of 611 PLHIV were included in the analysis to assess late HIV diagnostics using the LAg avidity, VL, and ARV detection algorithm. Overall, late HIV diagnosis was documented in 25.9% (95% CI: 21.7, 30.2) of adults in urban Ethiopia. The proportion of adults who tested for HIV late ranged from 38.1% in Gambela to 34.7% in Addis Ababa to 5.8% in Benishangul Gumuz to none in the Somali region (Table 3).

**Table 3:** Prevalence of late HIV diagnostics among people living with HIV (PLHIV) in urban-based population of Ethiopia

| Variables | Recent infection | Newly diagnosed | Prevalence of newly diagnosed |  |  |
| --- | --- | --- | --- | --- | --- |
|  | + self-reported | but long-term | but long-term infection |  |  |
|  | positive | infection | % | 95% CI |  |
| Region |  |  |  |  |  |
| Tigray | 35 | 4 | 10.3 | 4.2 | 23.1 |
| Afar | 24 | 8 | 24.9 | 14.4 | 39.6 |
| Amhara | 99 | 19 | 18.1 | 10.6 | 29.2 |
| Oromia | 108 | 41 | 31.0 | 22.8 | 40.5 |
| Somalia | 8 | 0 | 0.0 | 0.0 | 0.0 |
| Benishangul Gumuz | 19 | 1 | 5.8 | 1.0 | 26.7 |
| SNNPR | 32 | 17 | 34.7 | 20.7 | 51.9 |
| Gambella | 28 | 16 | 38.1 | 17.8 | 63.6 |
| Harari | 22 | 10 | 28.8 | 15.7 | 46.7 |
| Addis Ababa | 62 | 26 | 30.5 | 21.8 | 40.9 |
| Dire Dawa | 29 | 6 | 18.5 | 8.7 | 35.2 |
| Total | 464 | 147 | 25.9 | 21.7 | 30.5 |

As shown in Table 4, the proportion of late diagnosis of HIV among adult males was significantly higher, 36.8% (95%: 28.0, 46.6), than females, 20.8% (95%: 17.3, 24.9). The proportion of never married, married or living together PLHIV was slightly higher among those with late diagnosis, 30.3% and 29.2%, respectively, compared to the divorced, separated and widowed, 21.5% and 20.6%, respectively.

**Table 4:** Prevalence of late HIV diagnostics among people living with HIV (PLHIV) in urban-based population of Ethiopia,

| Variables | Recent infection | Newly diagnosed | Prevalence of newly diagnosed |  |  |
| --- | --- | --- | --- | --- | --- |
|  | + self-reported | but long-term | but long-term infection |  |  |
|  | positive | infection | % | 95% CI |  |
| Gender |  |  |  |  |  |
| Female | 363 | 98 | 20.8 | 17.3 | 24.9 |
| Male | 103 | 50 | 36.8 | 28.0 | 46.6 |
| Marital status |  |  |  |  |  |
| Never married | 49 | 22 | 30.3 | 19.9 | 43.3 |
| Married or living together | 212 | 73 | 29.2 | 22.3 | 37.3 |
| Divorced or separated | 114 | 30 | 21.5 | 14.6 | 30.6 |
| Widowed | 90 | 22 | 20.6 | 13.6 | 28.7 |
| Educational status |  |  |  |  |  |
| Not Educated | 98 | 23 | 16.4 | 10.7 | 24.4 |
| Primary | 217 | 74 | 28.4 | 22.4 | 35.3 |
| Secondary | 111 | 30 | 25.5 | 18.2 | 34.4 |
| More than secondary | 38 | 20 | 35.4 | 22.3 | 51.1 |
| Age |  |  |  |  |  |
| 15-24 | 35 | 27 | 43.5 | 28.6 | 59.6 |
| 25 -34 | 134 | 41 | 26.6 | 19.3 | 35.4 |
| 35 - 44 | 186 | 48 | 23.2 | 16.7 | 31.3 |
| 45 - 54 | 79 | 25 | 25.2 | 17.0 | 35.5 |
| 55 - 64 | 32 | 7 | 19.5 | 9.5 | 36.0 |
| Wealth quintile |  |  |  |  |  |
| Lowest | 77 | 26 | 31.7 | 20.8 | 45.0 |
| Second | 85 | 23 | 17.8 | 10.1 | 29.5 |
| Middle | 116 | 27 | 21.4 | 14.6 | 30.2 |
| Fourth | 107 | 40 | 29.0 | 20.4 | 39.4 |
| Highest | 81 | 32 | 30.5 | 21.5 | 41.3 |
| Drank Alcohol |  |  |  |  |  |
| Not reported drinking | 319 | 78 | 19.7 | 15.3 | 25.1 |
| Yes reported drinking | 147 | 70 | 36.1 | 29.3 | 43.5 |

Over one-third (35.4%) of the adult PLHIV attended secondary education and above were diagnosed late, followed by those with primary education (28.4%), secondary education (25.5%), and those who never attended school (16.4%). The highest proportion of late HIV diagnosis was observed among the adult PLHIV was in younger age group, 15 – 24 years (43.5%), followed by those 25–34 years (26.6%), and those 45–54 years (25.2%),. Late HIV diagnosis was the lowest among the 55– 64 year age group (19.5%) (Table 4).

PLHIV among adults with the lowest, fourth, and the highest quintiles had a higher proportion of late HIV diagnosis, 31.7%, 30, 5%, and 29.0%, respectively, while lower proportion was seen in second (17.8%) followed by the middle quintile (21.4%). The proportion of late diagnosis among PLHIV i who used alcohol was 36.1%, almost twice compared with adults not reported using alcohol (19.7%).

### Determinants for Never Tested for HIV

The results multilevel mixed-effect logistic regression model on diagnosis show that the model fitted well for the data over the standard logistic regression to assess determinants for never tested for HIV (X^2^=547.62, p<0.01). The result of ICC of the two-level multilevel model revealed that about 9.6% of the variation in the likelihood of never tested for HIV was explained by the variation among the regions in urban Ethiopia. As we can see from the 95% CI [0.043, 0.199], the variation among the regions is statistically significant and hence any estimation without considering this effect will result in a biased estimate (Table 5).

**Table 5:** Adjusted Odds Ratio (AOR) and 95% Confidence Interval (CI) for AOR of the multilevel mixed-effect logistic regression for the associated factors for never tested HIV in urban-based population of Ethiopia

| Variables | AOR | p-values | 95% CI for AOR |  |
| --- | --- | --- | --- | --- |
| Area |  |  |  |  |
| Small | 1.24 | 0.00 | 1.14 | 1.36 |
| Large | 1 |  |  |  |
| Gender |  |  |  |  |
| Female | 1 |  |  |  |
| Male | 1.39 | 0.00 | 1.29 | 1.50 |
| Marital status |  |  |  |  |
| Never married | 1 |  |  |  |
| Married or living together | 0.18 | 0.00 | 0.16 | 0.20 |
| Divorced or separated | 0.19 | 0.00 | 0.17 | 0.23 |
| Widowed | 0.20 | 0.00 | 0.16 | 0.25 |
| Educational status |  |  |  |  |
| Not Educated | 3.77 | 0.00 | 3.27 | 4.34 |
| Primary | 2.34 | 0.00 | 2.10 | 2.60 |
| Secondary | 1.62 | 0.00 | 1.45 | 1.79 |
| More than secondary | 1 |  |  |  |
| Age in years |  |  |  |  |
| 15-24 | 2.49 | 0.00 | 2.24 | 2.77 |
| 25 -34 | 1 |  |  |  |
| 35 - 44 | 1.54 | 0.00 | 1.35 | 1.75 |
| 45 - 54 | 3.14 | 0.00 | 2.72 | 3.64 |
| 55 - 64 | 5.26 | 0.00 | 4.48 | 6.17 |
| Wealth quintile |  |  |  |  |
| Lowest | 1 |  |  |  |
| Second | 0.73 | 0.00 | 0.65 | 0.82 |
| Middle | 0.67 | 0.00 | 0.59 | 0.75 |
| Fourth | 0.68 | 0.00 | 0.60 | 0.77 |
| Highest | 0.82 | 0.00 | 0.72 | 0.93 |
| Drank Alcohol |  |  |  |  |
| Not reported drinking | 1.37 | 0.00 | 1.26 | 1.48 |
| Yes reported drinking | 1 |  |  |  |
| Constant | 0.34 | 0.00 | 0.23 | 0.50 |
| Random intercept (Region) |  |  |  |  |
| Var(Uoi) | 0.35 |  | 0.15 | 0.82 |
| Region ICC | 0.096 |  | 0.043 | 0.199 |
| LR test vs. logistic model: $\text{chibar2}(01) = 547.62$ Prob $\geq \text{chibar2} = 0.0000$ | | | | |

Adults aged 15–24 years (AOR: 2.49; 95% CI: 2.24, 2.77), 35–44 years (AOR: 1.54; 95% CI: 1.35, 1.75), 45–54 years (AOR: 3.14; 95% CI: 2.72, 3.64), and those 55–64 years (AOR: 5.26; 95% CI: 4.48, 6.17) were 2.49, 1.54, 3.14, and 5.26 times more likely to have never tested for HIV, respectively, as compared to those 25–34 years of age. The odds of being never tested for HIV among adults living in small urban areas were 1.24 times higher than those living in large urban areas (AOR: 1.24; 95% CI: 1.14, 1. 36). Being male was significantly and positively associated with never tested status compared with the female counterparts (AOR: 1.39; 95% CI: 1.29, 1.50) (Table 5).

Adults who were married or living together, divorced or separated, and widowed had 82.0% (AOR: 0.18; 95% CI: 0.16, 0.20), 81.0% (AOR: 0.19; 95% CI: 0.17, 0.23), and 80.0% (AOR: 0.20; 95% CI: 0.16, 0.25), respectively, lower odds of being never tested for HIV compared to their never married counterparts. In terms of education level of respondents who never attended school (AOR: 3.77; 95% CI: 3.27, 4.34), primary (AOR: 2.34; 95% CI: 2.10, 1.79), and secondary education (AOR: 1.62; 95% CI: 1.45, 1.79) were 3.77, 2.34 and 1.62 times more likely to never tested for HIV, respectively as compared to those who had secondary education or above (Table 5).

Adults who were in second, middle, fourth, and highest wealth quintiles had 27.0% (AOR: 0.73; 95% CI: 0.65, 0.82), 33.3% (AOR: 0.67; 95% CI: 0.59, 0.75), 32.0% (AOR: 0.68; 95% CI: 0.60, 0.77), and 18.0% (AOR: 0.82; 95% CI: 0.72, 0.93), respectively, lower odds of being never tested compared to their counterparts in the lowest quintiles. The odds of being never tested for HIV among adults aged 15 to 64 who had not drink alcohol (AOR 1.37; 95% CI; 1.26, 1.48) was 1.37 times higher compared to those who used alcohol.

### Determinants for Late Diagnostics for HIV

The results on the diagnosis of multilevel mixed-effect logistic regression model showed that the model fitted well for the data over the standard logistic regression to assess determinants for late diagnosis for HIV (X2=6.6, p<0.01). The result of ICC of the two-level multilevel model revealed that about 6% of the variation in the likelihood of never tested for HIV was explained by the variation among the regions in urban Ethiopia, As seen from the 95% CI [0.012, 0.257], the variation among the regions is statistically significant and hence any estimation without considering this effect will result in a biased estimate (Table 6).

**Table 6:** AOR and 95% CI for AOR of the multilevel mixed-effect logistic regression for the associated factors for lately tested HIV among PLHIV in urban-based population of Ethiopia

| Variables | AOR | p-value | 95% CI for AOR |  |
| --- | --- | --- | --- | --- |
| Gender |  |  |  |  |
| Female | 1 |  |  |  |
| Male | 1.69 | 0.02 | 1.09 | 2.63 |
| Age |  |  |  |  |
| 15-24 | 1 |  |  |  |
| 25 -34 | 0.41 | 0.01 | 0.22 | 0.79 |
| 35 - 44 | 0.32 | 0.00 | 0.17 | 0.60 |
| 45 - 54 | 0.34 | 0.00 | 0.17 | 0.70 |
| 55 - 64 | 0.24 | 0.01 | 0.09 | 0.66 |
| Drank Alcohol |  |  |  |  |
| Not reported drinking | 1 |  |  |  |
| Yes reported drinking | 2.07 | 0.00 | 1.38 | 3.09 |
| Constant | 0.47 | 0.02 | 0.25 | 0.90 |
| Random intercept (Region) |  |  |  |  |
| Var(Uoi) | 0.21 |  | 0.04 | 1.14 |
| Region ICC | 0.06 |  | 0.012 | 0.257 |
| LR test vs. logistic model: $\text{chibar2}(01) = 6.55$ | | Prob $\geq \text{chibar2} = 0.0052$ | | |

Among PLHIV aged 15 to 64 years, being male was 1.69 times more likely to present late for HIV diagnosis as compared to those female counterparts within the regions (AOR: 1.69; 95% CI: 1.09, 2.63). PLHIV in the age group of 25–34 years, 35–44 years, 45–54 years, and those 55–64 years had 59% (AOR: 0.41; 95% CI: 0.22, 0.79), 68.0% (AOR: 0.32; 95% CI: 0.17, 0.60), 66.0% (AOR: 0.34; 95% CI: 0.17, 0.70), and 76.0% (AOR: 0.24; 95% CI: 0.09, 0.66), respectively, lower odds of being lately diagnosed for HIV compared to those 15–24 years of age with in the regions (Table 6).

The odds of late diagnosis for HIV among PLHIV aged 15 to 64 who ever had used alcohol (AOR 2.07; 95% CI; 1.38, 3.09) was 2.07 times higher compared to those who had drink with in the regions.

## Discussion

As the gateway to a continuum of HIV/AIDS services, timely HIV testing plays a central role in the fight against the HIV epidemic [22]. A recent study demonstrated the efficacy of early testing in immediately keeping patients on ART for the prevention of HIV transmission; and studies based on mathematical models suggest that the Test and Treat strategy, consisting of treating every HIV-infected person as soon as diagnosis is made, can curb the epidemic [2,3]. For all these reasons, there is currently an international consensus to expand HIV testing in resource limited countries like Ethiopia, as part of which generating evidence in this field will be crucial.

### HIV testing uptake and its determinants

In this analysis 29.4% of adults in urban Ethiopia were never tested for HIV. Compared to the national standard of 90% testing in the general population, this study revealed that the uptake of HIV testing in urban Ethiopia is still low. It was consistent with a study conducted in sub-Saharan Africa that revealed 25% of population in Malawi reported having ever been tested for HIV. Similar study reported lower HIV testing uptake in Côte d’Ivoire (50%) and Mozambique (51%) [1].

Furthermore, our finding revealed that HIV testing uptake differed across the administrative regions in in Ethiopia. The proportion of adults who reported being never tested for HIV ranged from 18.8% in the Harari region to 20.3% in Afar to 32.6% in Oromia and to 69.1% in Somalia. This pattern was significantly different across regions with non-overlapping 95% CIs. This suggests that programmatic interventions for HIV testing may need to be made based on the context of regions.

Our study showed that the proportion of never tested among adult males was significantly higher than females. This is consistent with evidence from Zimbabwe, which showed a significant difference between male and female testing uptake [16]. In contrast, a study conducted in Uganda reported a lower proportion of males (16%) than females was never tested [24]. Other studies have shown that women are more likely to report having ever been tested than men [5, 4, 7, 8]. In our study, among adults aged 15 to 64 years being male was significantly associated with never tested for HIV compared with the female counterparts in the regions. This might be explained by the high rate of testing uptake for female as a result of PMTCT during antenatal follow-up.

Our study also reveals that there were significant variations in being never tested by education status. The proportion of never tested for HIV was higher among adults with no education, primary education or secondary education, 33.5%, 32.3%, 32.4%, respectively, compared to those with more than secondary school education (19.8%). This was similar with the finding of a study conducted in Ghana, which revealed the increased testing uptake as the level of education increases, ranging from 14.2% to 71.1% [27], while there was no significant difference in by education categories in a study conducted in Cote d’Ivoire, which ranged from 65% to 69.6% [22].

Similarly, our study showed that the proportion of adults never tested was highest at age 15–24 years (47.8%) followed by 55–64 years (41.2%). Adults aged between 15 -24, 35–44, 45–54 and 55–64 years were 2.49, 1.54, 3.14, and 5.26 times more likely to have never tested for HIV compared to 25–34 years, respectively. This pattern across age categories was significantly different, 95% CIs non-overlapping. Adults in the age group of 15–24 years (AOR: 2.49; 95% CI: 2.24, 2.77), 35–44 years (AOR: 1.54; 95% CI: 1.35, 1.75), 45–54 years (AOR: 3.14; 95% CI: 2.72, 3.64) and those 55–64 years (AOR: 5.26; 95% CI: 4.48, 6.17) were 2.49, 1.54, 3.14, and 5.26 times more likely to being never tested for HIV, respectively, as compared to those 25 – 34 years of age in the regions. This may highlight the need for the program to focus on addressing specific age groups, including in promotion of school and enhanced elderly testing strategy since HIV incidence in the age 15-24 years is the highest among all age group (0.12) and HIV prevalence among the elderly accounts for 4.4%, compared to the prevalence in the general population of 0.96%[10, 11].

Adults who were married or living together, divorced or separated, and widowed had 82.0% (AOR: 0.18; 95% CI: 0.16, 0.20), 81.0% (AOR: 0.19; 95% CI: 0.17, 0.23), and 80.0% (AOR: 0.20; 95% CI: 0.16, 0.25), respectively lower odds of being never tested for HIV compared to their never married counterparts in the regions. In terms of education level of respondents who had never attended school (AOR: 3.77; 95% CI: 3.27, 4.34), had primary education (AOR: 2.34; 95% CI: 2.10, 1.79), and had secondary education (AOR: 1.62; 95% CI: 1.45, 1.79) were 3.77, 2.34 and 1.62 times more likely to never tested for HIV, respectively as compared to those who had above secondary education with in the regions. This finding is peculiar to Ethiopia since other reports from Uganda, Ghana and Zimbabwe showed no difference among these sub-population groups [28][24][7]. This, in the context of Ethiopia, could be worrisome for the program since the prevalence of HIV among the widowed is significantly higher 14.0% compared to 0,96% in the general and 3.0% in urban Ethiopia [11,10].

In terms of wealth quartile, 39.3% of the adults who were in the lowest quartile were never tested for HIV, significantly more respondents who were in the second, third, fourth, and highest quartiles (30.0%, 26.3%, 26.6%, and 27.2%, respectively) with non-overlapping 95% CIs. However, this finding was inconsistent with a study conducted in Uganda and South Africa [12,6]. Similarly, the proportion of those never tested for HIV among adults who were not employed (during the last 12 months) preceding the survey was 37.0%, significantly more than (almost twice) those employed (21.0%), 95% CIs non-overlapping. This is in line with the global recommendations towards income generating activities as part of HIV prevention [14, 15].

### Late HIV diagnosis and its determinants

By assessing late HIV diagnostics using the LAg avidity, VL, and ARV detection algorithm, overall, 25.9% (95% CI: 21.7, 30.2) of the adults were diagnosed late for HIV in urban Ethiopia. This was high compared to the finding of a study conducted in united kingdom (UK), which showed that 15.4% were diagnosed late [28], while our study is consistent with a report from Spain, which showed 30.4% were late presenters [18].

In terms of regions, the proportion of adults who tested late for HIV ranged from 38.1% in the Gambela region to 34.7% in Addis Ababa to 5.8% in Benishangul Gumuz and to 0.0% in Somalia. Given the heterogeneity of HIV epidemic prevalence by region in Ethiopia, which ranged from 5.6% in Gambella followed by 3.3% in Addis Ababa and is less than 1% in Somali [9] and that the highest proportion of people with late diagnosis is in the high burden regions, the program may consider focusing on devising strategies to increase testing uptake in the high burden regions.

As shown in our study, the proportion of late diagnosis for HIV among adult males was significantly higher (36.8%; 95%: 28.0, 46.6) than females (20.8%; 95%: 17.3, 24.9). This result was not in line with a study in Mozambique which reported late diagnosis among males Vs females of 43.7% and 55.3%, respectively. However, our finding concurs with the findings of a study from South Africa which revealed that 57% of males and 43% of females were diagnosed late [30], showing that the proportion late diagnosis is higher among males.

Among PLHIV aged 15 to 64 years, males were 1.69 times more likely to present late for HIV diagnosis as compared to their female counterparts in the regions (AOR: 1.69; 95% CI: 1.09, 2.63). Likewise, PLHIV in the age group of 25–34 years, 35–44 years, 45–54 years and those 55–64 years had 59.0% (AOR: 0.41; 95% CI: 0.22, 0.79), 68.0% (AOR: 0.32; 95% CI: 0.17, 0.60), 66.0% (AOR: 0.34; 95% CI: 0.17, 0.70), and 76.0% (AOR: 0.24; 95% CI: 0.09, 0.66), respectively, and had a lower odds of being diagnosed late for HIV compared to those 15–24 years of age. In-terms of age distribution, the highest proportion of late diagnosis for HIV among adults living with HIV was in younger age 15–24 years (43.5%), followed by 25–34 years (26.6%) and in 45–54 years (25.2%), the lowest was in 55–64 years (19.5%). This may indicate the need for a more rigorous programmatic intervention among the adolescent age group to enhance early diagnosis.

A multilevel mixed-effect logistic regression model showed that the model fitted well for the data over the standard logistic regression to assess determinants of late HIV diagnosis (*X*^*2*^=6.6, p<0.01). The result of ICC of the two-level multilevel model revealed that about 6% of the variation in the likelihood of never tested for HIV was explained by the variation among the regions in urban Ethiopia As we can see from the 95% CI: 0.012, 0.257, the variation among the regions is statistically significant and hence any estimation without considering this effect will result in a biased result.

## Conclusion and recommendation

The country is still far behind the global target of 90% with regard to HIV testing uptake. HIV testing offer should be focused on geographically highly affected regions which may include Gambella and Addis Ababa. The significant low male testing uptake may need innovative approach to address. The low coverage of young adolescent testing may call up the establishment to a school testing strategy. Moreover, the highly affected widowed population with low level of testing uptake shall be a focus area for the program. There shall be a strategy to address disadvantaged populations, especially those with limited education and low income.

Late-stage HIV testing has diverse domains of barriers, including demographic, economic, geographic and social factors. Efforts should focus on various interventions, such as multispectral engagement, community engagement, income generating activities and the use of active mobile testing strategies to reduce late-stage disease presentation. Providing increased and targeted HIV testing to those at greatest risk for late-stage HIV disease and subsequently linking them to HIV care should be a priority so that to reduce AIDS-related morbidity and mortality.

### Strength and Limitation of the study

This study used a huge data set and has a potential to represent the urban population in Ethiopia. However, more than 85% of the population Ethiopia lives in rural area and this study didn’t address this population living in the rural area.

## Data statement

The data source for this study is publicly available as part of the Ethiopian Population Based HIV impact Assessment.

## Data Availability

https://www.aarc.gov.et/index.php/resources/and-on-health-related-issues/ethiopia-data-information-on-hiv-aids

## Funding

Since this study used a secondary data from the Population Based HIV impact assessment, there was no funding for this study.

## Competing interest

Authors declare there is no competing interest for the study.

## Acknowledgement

We would like to acknowledge the Ethiopian Public Health Institute that led this study and the collaborating partners including CDC and ICAP Ethiopia. Last but not list, would like to acknowledge study participants and the data collection team.

## Author statement

Yimam Getaneh conducted this study since the inception to the write-up while Jemal Ayalew supported on data analysis and write-up. Ebba Abate and Adamu Tayachew reviewed the study and support during the analysis of the study. Sileshi Lulseged and Yiming Shao oversee the study since the inception to the write-up of the manuscript.

## Notes

### Competing Interest Statement

The authors have declared no competing interest.

### Author Declarations

Ethical considerations The study protocol obtained ethical clearance from the institution review boards (IRBs) of the Ethiopian Public health Institute, Centers for Disease Control and Prevention (Atlanta), and Columbia University (New York). The EPHIA Data Analysis Advisory Committee at EPHI approved the data analysis. Study participants gave consent for this study in addition to the IRB approvals.

